# Acute and chronic psychosocial stress by the brain-derived neurotrophic factor in male humans: a highly standardized and controlled study

**DOI:** 10.1101/2023.09.29.23296327

**Authors:** Benedict Herhaus, Martin Heni, Wilhelm Bloch, Katja Petrowski

## Abstract

**Objective:** The neurotrophic protein brain-derived neurotrophic factor (BDNF) plays a pivotal role in brain function and is affected by acute and chronic stress. We here investigate the patterns of BDNF and cortisol stress reactivity and recovery under the standardized stress protocol of the TSST and the effect of perceived chronic stress on the basal BDNF levels in healthy young men.

**Methods:** Twenty-nine lean young men underwent the Trier Social Stress Test (TSST) and a resting condition. Serum BDNF and cortisol were measured before and repeatedly after both conditions. The perception of chronic stress was assessed by the Trier Inventory for Chronic Stress (TICS).

**Results:** After the TSST, there was a significant increase over time for BDNF and cortisol. Stronger increase in cortisol in response to stress was linked to an accelerated BDNF decline after stress. Basal resting levels of BDNF was significantly predicted by chronic stress perception.

**Conclusions:** The increased BDNF level following psychosocial stress suggest a stress-induced neuroprotective mechanism. The presumed interplay between BDNF and the HPA-axis indicates an antagonistic relationship of cortisol on BDNF recovery post-stress. Chronically elevated high cortisol levels, as present in chronic stress, could thereby contribute to reduced neurogenesis, and an increased risk of neurodegenerative conditions in persons suffering from chronic stress.

**Highlights:**

- Acute psychosocial stress increases serum BDNF and cortisol
- Stress-induced cortisol secretion may accelerate the decline of BDNF after stress.
- Chronic stress is linked to lower basal serum BDNF levels

## 1. Introduction

Stress-related disorders have emerged as a major health concern in the early twenty-first century and have been linked to altered brain function [1,2]. Acute and chronic stress affects adult neurogenesis and activates a complex interplay of neural and endocrine mechanisms [3,4]. Glucocorticoids (GCs), as a key part of the body’s stress response system, and brain-derived neurotrophic factor (BDNF), with its multiple roles in the nervous system, influence adult neurogenesis through their dynamic interactions in the context of acute and chronic stress [5,6].

The neurotrophic protein BDNF plays a pivotal role in brain function throughout life [6]. BDNF protects existing neurons and synapses of the central nervous system (CNS) [7]. In the neurogenesis BDNF acts through the family of high-affinity tyrosine kinase receptors i.e. the tropomyosin receptor kinase B (TrkB) receptor [8,9]. In addition to these direct functions on neuronal structure, BDNF is known to contribute to the regulation of eating behaviour and physical activity [9]. In the hippocampal, BDNF is a key molecule related to learning and memory [6]. BDNF physiologically decreases with normal ageing [10] and the development of cognitive impairment is affected by chronic stress [11]. Of note, altered BDNF were detected in different diseases, including depression [12] and Alzheimer’s disease [13].

Different studies suggested that both acute and chronic stress could affect BDNF [6]. With regard to the acute stress, BDNF increases have been observed in healthy adults in response to acute psychosocial stress in the laboratory using the internationally established Trier Social Stress Test [14–16]. In animals, chronic stress caused a reduction in neuronal *Bdnf* mRNA expression [17]. In line, reduces BDNF levels were reported in hospital employees who were psychologically stressed through their work [18]. Thus, BDNF is a stress-dependent factor [19].

Of note, BDNF appears to interact with the hypothalamic-pituitary-adrenocortical (HPA)-axis, one of the major physiological stress systems. *BDNF* is expressed in the hypothalamus’s paraventricular nucleus (PVN) [20], the upstream regulator of HPA-axis activity. In this brain area, corticotrophin-releasing hormone (CRH) is regulated in response to internal and external stimuli [21]. Glucocorticoid action on glucocorticoid receptors in the PVN are crucial components of the negative feedback loop that controls HPA-axis activity [22]. Of notice, dysregulation of glucocorticoid receptor function not only leads to a disturbed HPA-axis feedback loop with an increase in CRH-expression but also causes an increase of hypothalamic BDNF [19]. This suggests a causal suppressive impact of glucocorticoid signaling on BDNF expression or release. Though, the relation between glucocorticoids and BDNF appears to be bi-directional, as BDNF injections also raised CRH levels and stimulated the HPA-axis [23]. How these mechanistic findings translate into humans is still not fully clear.

The HPA-axis regulates the acute and chronic stress response through the secretion of glucocorticoids, the most important of which is cortisol [5]. While it is well established that BDNF and cortisol levels increase in response to acute stress [6], how they interact under acute stress is less clear. As a possible mechanism, Suri & Vaidya [24] described that stress-induced cortisol secretion has regulatory effect on BDNF secretion.

Human studies have shown stress-induced increases in both cortisol and BDNF in response to a psychosocial stressor [14–16]. However, possible interactions between BDNF and cortisol dynamics are not well-established. The three previous studies on the topic had varied methodologies. Factors like a single post-stress blood sample [14,15], salivary cortisol measurements [15], and no distinct analysis of BDNF/cortisol responses to stress and recovery [14–16] make it hard to definitively determine their relationship.

Therefore, we aimed to clarify the patterns of BDNF and cortisol stress reactivity and -recovery under the standardized stress protocol of the TSST with repeated blood samples to determine cortisol and BDNF in parallel. Based on the previous studies investigating BDNF and cortisol stress reactivity [14–16], we hypothesized stress-induced increase of BDNF and cortisol following the acute psychosocial laboratory stressor (Hypothesis 1). Given the postulated regulatory effect of stress-induced cortisol on BDNF secretion [24], we hypothesized a negative association between patterns of cortisol stress reactivity and BDNF stress recovery (Hypothesis 2). In view of the relationship between chronic stress and BDNF levels [17,18], we hypothesized that chronic stress perception would be a predictor of basal resting levels of BDNF (Hypothesis 3).

## 2. Methods

### 2.1 Study participants

Twenty-nine healthy male individuals were recruited via electronic tendering (e-tendering) and notice boards at the Johannes Gutenberg University Mainz. Study criteria were assessed by telephone interview using the full Structured Clinical Interview (SCID; [25]) for the Diagnostic and Statistical Manual of Mental Disorders (DSM-IV, [26]). Exclusion criteria were acute or chronic medical illness, mental disorders, medication or substance use, stressful life events in the past six months, being younger than 18 or older than 35, and smoking more than ten cigarettes per day. Due to known age and gender specific differences in BDNF levels [27,28], only male participants aged between 18 and 35 years were included. The mean age of the participants was 24.34 ± 4.08 years of age with a body mass index (BMI) of 22.94 ± 1.61 kg/m². A detailed description of all participants, including demographic data and psychological status is given in Table 1. The study protocol was approved by the local Ethics Committee of the Landesärztekammer Rheinland-Pfalz, Germany (No#2019-14188).

**Table 1.** Characteristics of the male participants.

|  | Individuals (N=29) |
| --- | --- |
| *Demographic data* |  |
| Age (years) | 24 (4) |
| Body mass index (kg/m<sup>2</sup>) | 22.9 (1.6) |
| Smoking, n (%) | 3 (10) |
| *Psychological Assessment* |  |
| BDI | 5.62 (4.67) |
| SCL Global Severity Index | .36 (.29) |
| PSS | 22.24 (6.78) |
| TICS-SCSS | 12.10 (6.26) |

Data are presented as mean (standard deviation)
*BDI* Beck Depression Inventory, *PSS* Perceived Stress Scale, *SCL* Symptom-Check-List-90-R, *SCSS* Subscale of Chronic Stress, *TICS* Trier Inventory of Chronic Stress.

**Table 2.** BDNF, cortisol, and subjective appraisal during rest and in response to the Trier Social Stress Test.

| **Healthy Men (N=29)** |  |  |  |  |
| --- | --- | --- | --- | --- |
|  | Resting Condition | TSST | Dependent Student *t*-test |  |
| **Derived BDNF parameters (pg/ml)** | M (SD) | M (SD) | *t* | *p* |
| Peak | 33565 (10566) | 37677 (10881) | 4.536 | $\leq .001$<br>(*d* = .84) |
| Delta Peak-Baseline | 4264 (5459) | 7673 (5488) | 2.514 | $\leq .01$<br>(*d* = .47) |
| AUC<sub>I</sub> | 21881 (126123) | 99503 (104316) | 2.555 | $\leq .01^{**}$<br>(*d* = .47) |
| **Derived cortisol parameters (ng/ml)** |  |  |  |  |
| Peak | 57.31 (15.88) | 92.20 (24.76) | 7.831 | $\leq .001$<br>(*d* = 1.47) |
| Delta Peak-Baseline | 4.96 (8.37) | 40.08 (27.31) | 6.736 | $\leq .001$<br>(*d* = 1.27) |
| AUC<sub>I</sub> | -90.23 (273.66) | 757.66 (689.28) | 5.870 | $\leq .001$<br>(*d* = 1.11) |
| **Subjective Appraisal** |  |  |  |  |
| PASA - Stress index | -2.68 (1.28) | -0.54 (1.39) | 7.663 | $\leq .001$<br>(*d* = -1.41) |
| VAS | 41.35 (11.23) | 58.51 (9.42) | 8.903 | $\leq .001$<br>(*d* = -1.65) |

*AUC<sub>I</sub>* incremental area under the curve with respect to increase, *M* Mean, *PASA* Primary Appraisal Secondary Appraisal, *SD* Standard Deviation, *VAS* Visual Analogue Scale.

### 2.2. Study design

Participants underwent two different conditions, stress and resting, on separate days within a seven-day period. Both conditions started between 2:00 p.m. and 5:00 p.m., and the order of testing was randomized. Participants were asked to refrain from eating, drinking, and smoking before and during the two-hour test session. To avoid a pain-induced release of BDNF and cortisol, the intravenous cannula was inserted 45 minutes before the first blood sample was taken. The experimental protocol began with a 15-minute pre-session, which included the collection of two blood samples. Participants then underwent two 15-minute conditions: stress and rest. The stress condition was the Trier Social Stress Test (TSST), following the Kirschbaum et al. [29] protocol. It consisted of three sections: preparation, interview, and a calculation task, with each section lasting 5 minutes. In contrast, during the resting condition, participants were given the opportunity to read magazines. Cognitive appraisal was assessed three minutes after the start of each condition using the Primary Appraisal Secondary Appraisal (PASA) scale [30]. In addition, participants’ self-reported perception of stress was measured immediately after both conditions using the visual analogue scale (VAS). After completing the stress and resting conditions, participants remained in a supine position on a bed for 105 min, during which nine blood samples were taken at different time points: +1, +5, +10, +20, +30, +45, +60, +75, and +105 minutes.

### 2.3 Blood analytics

Blood samples were collected in serum monovettes (S-Monovette® 7.5 ml Z, Sarstedt, Nümbrecht, Germany). After blood collection, the monovettes were left at room temperature for 30 min to allow the blood to coagulate. The monovettes were then centrifuged at 2500 g for 10 min at 20°C, divided into aliquots and stored at -80°C. BDNF concentration were determined by enzyme-linked immunosorbent assay (ELISA) (Human Free BDNF Quantikine ELISA Kit– R&D Systems Europe, Ltd. Abingdon, United Kingdom). Serum cortisol concentrations were quantified using a commercially available enzyme-linked immunosorbent assay (ELISA) kit (IBL International GmbH, Germany).

### 2.4 Questionnaires

The Beck Depression Inventory (BDI; [31]) was used to assess the severity of depression. The inventory is based on 21 items with a four-point rating scale from 0 to 3. The total score ranges from 0 and 63, with a higher total score indicating more depressive symptoms. The Perceived Stress Scale (PSS; [32]) measures the level of stressful situations in in one’s life during the previous month. The questionnaire consists of 14 items on a 5-point scale ranging from 1 ‘never’ to 5 ‘very often’. The Global Severity Index (GSI) of the Symptom Checklist-90- Revised (SCL-90-R; [33]) was used to assess a persońs perceived impairment from physical and psychological symptoms of a person. The screening subscale (SCSS) of the Trier Inventory for Chronic Stress (TICS), developed by Schulz et al. [34], was used to assess the level of chronic stress experienced in the previous three months. Eleven items have to be answered on a five-point rating scale ranging from ‘never’ (0) to ‘very often’ (4).

### 2.5 Statistical analysis

A power analysis calculated with the G*power program (version: 3.1.9.2.) [35] showed that to expect a medium effect size of Cohen’s f =.25 for the outcome measure of BDNF, using a two-way MIXED ANOVA for repeated measures as the statistical test to prove interaction of within-factor time (measurement points -1, +1, +5, +10, +20) and within-factor condition (stress vs. resting) with a significance level of p =.05 and power of 80 % (1-β =.80), a total sample size of n = 22 participants would be required. The BDNF and cortisol data were analyzed according to the normality of the distributions and, in case of non-normally distributed data, were subjected to logarithm naturalis transformations. Statistical analysis was performed with SPSS Statistics version 27 (IBM, Chicago, IL, USA).

For the BDNF and cortisol response, the area under the curve with respect to increase (AUCI) and the delta between peak and baseline (Δ Peak-Base) were calculated [36,37]. All parameters (PASA, VAS, Cortisol-AUCI, & Cortisol Δ Peak-Base) were analyzed by two-factorial MIXED ANOVA for repeated measurements with the within-factor condition (stress vs. resting) and within-factor time.

For the specification of the BDNF and cortisol stress reactivity and recovery during the stress condition, the area under the curve with respect to increase and decrease (AUCI /AUCD) were calculated using the formulas of Pruessner et al. [36]. Stress reactivity was defined as the incremental area under the curve from baseline to peak value. Stress recovery was defined as the decremental area under the curve from the peak value to the last measurement point. Both areas under the curve were calculated individually for each subject based on the individual peak values. The association between BNDF and cortisol stress reactivity and recovery was tested using Pearson’s correlation test.

Regression was calculated to predict the influence of the subjective chronic stress on the basal serum BNDF concentration (-1 minute time point) of the resting and stress condition.

## 3. Results

*BDNF responses to acute stress compared to rest* BDNF levels were comparable before both the stress and the resting condition (-1min: *t* (28) =.433, *p* =.67, Figure 1). ANOVA results indicated a significant effect of time over the five measurement points (*F* (2.922, 81.820) = 8.110, *p* ≤.001, *η2* =.225). There was a significant main effect of condition on BDNF concentration, with higher values in the stress induction condition compared to the resting condition (*F* (1, 28) = 6.506, *p* ≤.05, *η2* =.189) and a significant interaction effect time x condition (*F* (4, 112) = 5.532, *p* ≤.001, *η2* =.165). In line, peak concentrations in BDNF (*t* (28) = 4.536, *p* ≤.001, *d* =.84), the absolute change in BDNF (*t* (28) = 2.514, *p* ≤.01, *d* =.47), and the incremental AUCI (*t* (28) = 2.555, *p* ≤.01, *d* = -.47) were higher in the stress condition compared to the resting condition.

**Figure 1.**
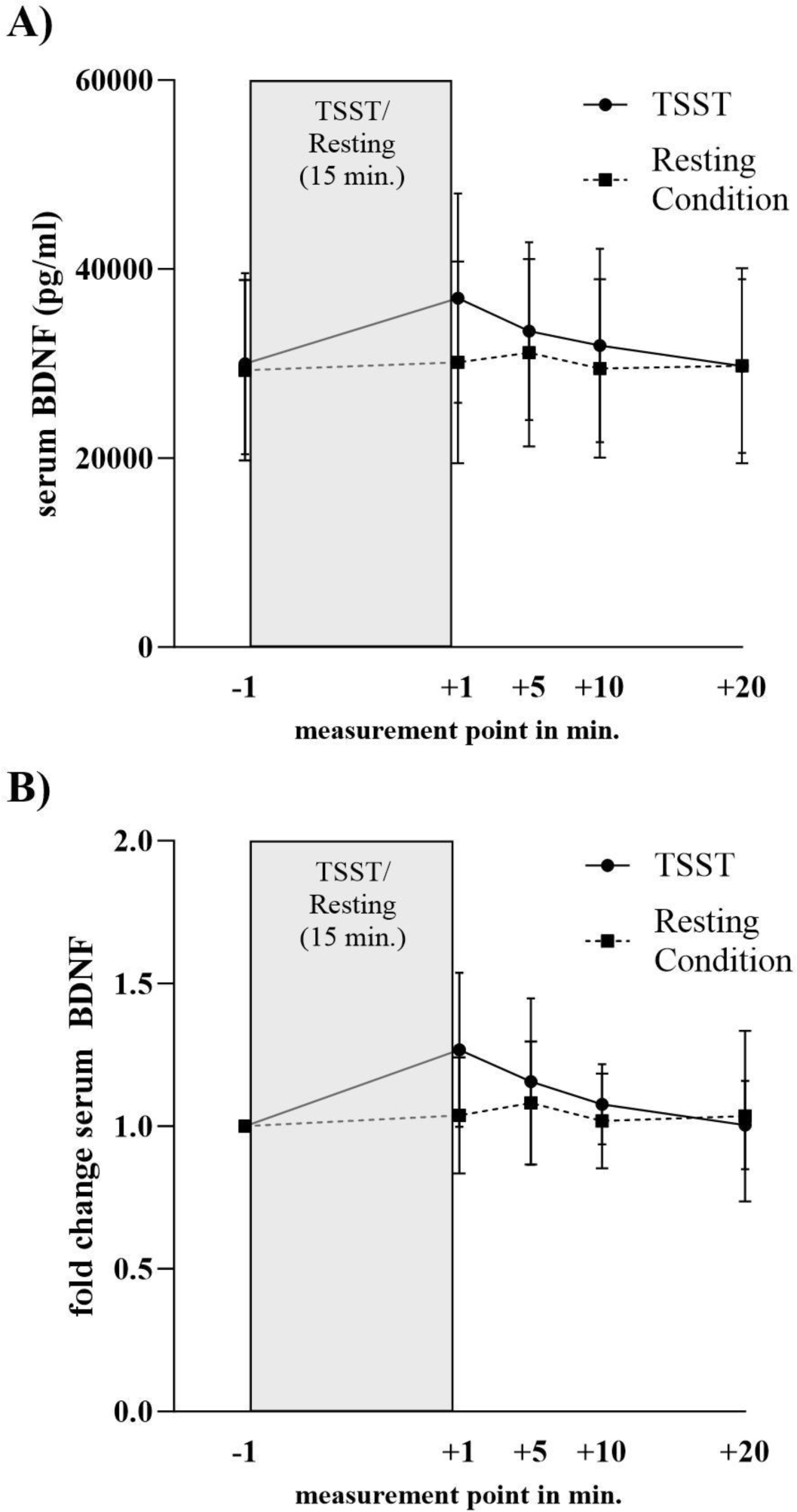
BDNF concentration during Trier Social Stress Test and resting condition.Presented are means +/- SD in healthy men (N=29).

### Cortisol responses to acute stress compared to rest

Resting serum cortisol was comparable between the two study days (-1min: *t* (27) = -.089, *p* =.93, Figure 2). There were significant time (*F* (3.691, 99.653) = 63.186, *p* ≤.001, *η2* =.701), condition (*F* (1, 27) = 40.111, *p* ≤.001, *η2* =.598), and time x condition effect (*F* (4.125, 111.386) = 16.947, *p* ≤.001, *η2* =.386) on serum cortisol. Peak cortisol (*t* (27) = 7.831, *p* ≤.001, *d* = 1.47), the absolute change in cortisol (*t* (27) = 6.736, *p* ≤.001, *d* = 1.27), as well as the incremental AUCI (*t* (27) = 5.870, *p* ≤.001, *d* = 1.11) were higher in the stress condition compared to the resting condition.

**Figure 2.**
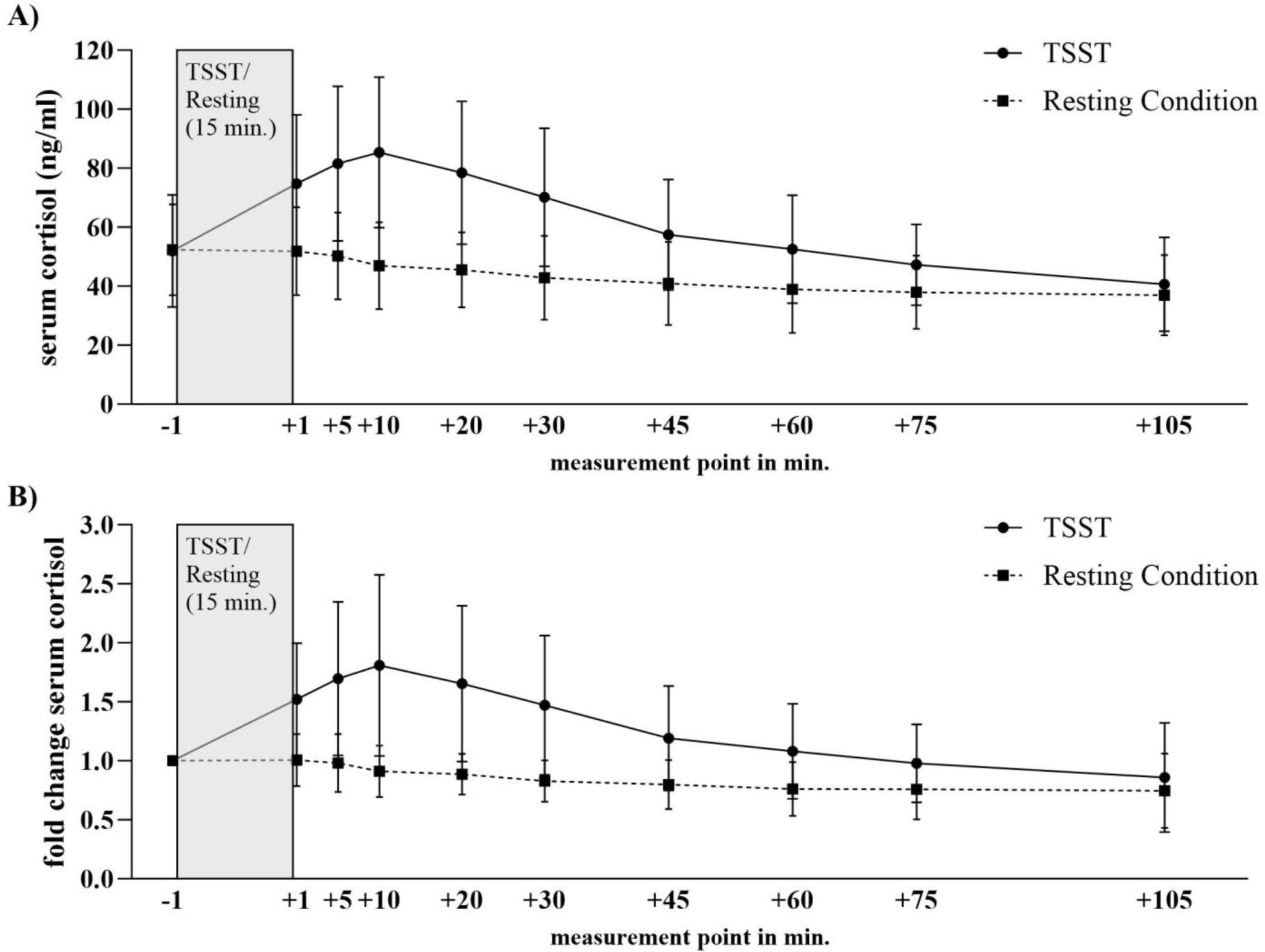
Cortisol concentration during Trier Social Stress Test and resting condition. Presented are means +/- SD in healthy men (N=29).

### Psychological responses to stress induction versus rest

Analyses of the two questionnaires showed that the TSST was successful in inducing stress: male participants showed significantly higher scores in the VAS (t (28) = 7.633, p ≤.001, d = 1.41) and on the tertiary scale ‘stress index‘ of the PASA (*t* (28) = 8.903, *p* ≤.001, *d* = 1.65).

### Association between BDNF and cortisol stress reactivity and their recoveries

Regarding to the relationship between BDNF and cortisol reactivity/recovery patterns, a negative significant correlation was found between cortisol stress reactivity and BDNF stress recovery (*r* (26) = -.39, *p* ≤.05). No significant correlation was found between BDNF and cortisol stress recovery nor between BDNF stress reactivity and cortisol stress reactivity/recovery (see Table 3).

**Table 3.**
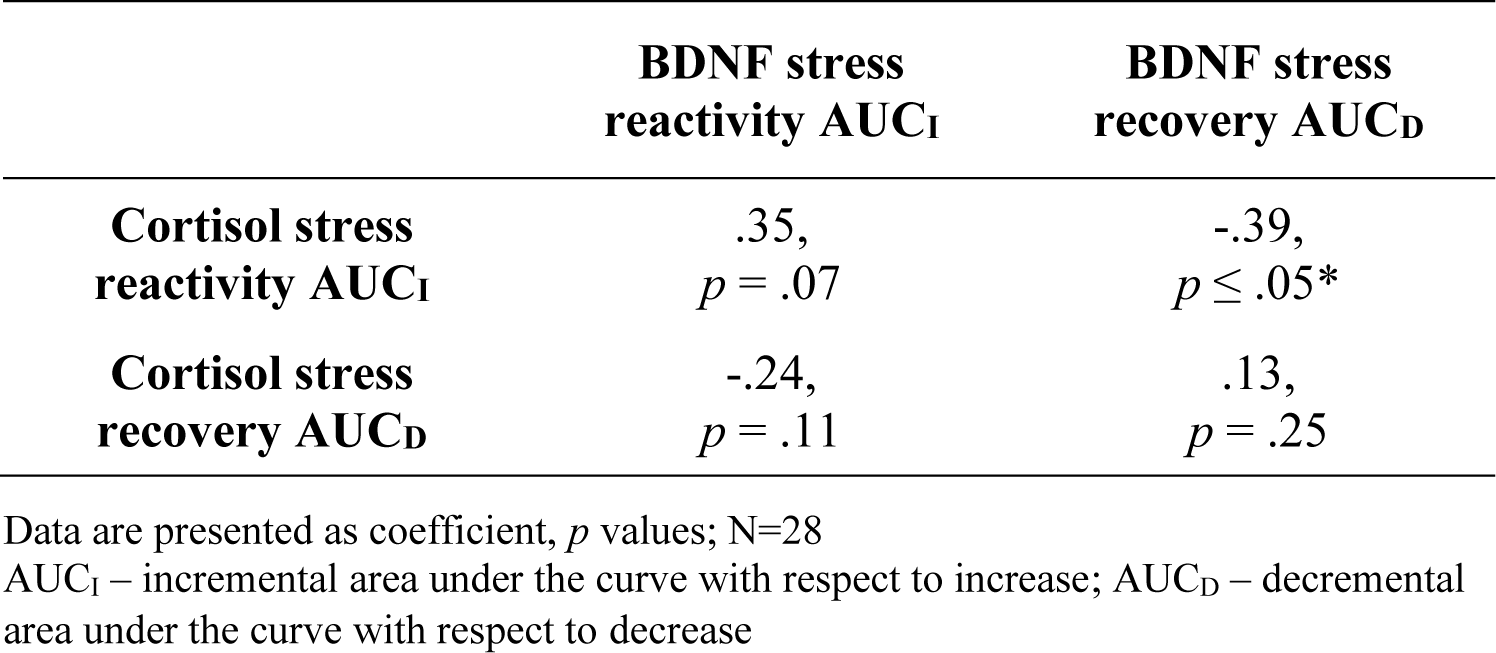
Pearson’s correlations (r) between BNDF and cortisol stress reactivity and stress recovery.

|  | **BDNF stress**<br>**reactivity AUC<sub>I</sub>** | **BDNF stress**<br>**recovery AUC<sub>D</sub>** |
| --- | --- | --- |
| **Cortisol stress**<br>**reactivity AUC<sub>I</sub>** | .35,<br>*p* = .07 | -.39,<br>*p* ≤ .05* |
| **Cortisol stress**<br>**recovery AUC<sub>D</sub>** | -.24,<br>*p* = .11 | .13,<br>*p* = .25 |

Data are presented as coefficient, *p* values; N=28
AUC<sub>I</sub> – incremental area under the curve with respect to increase; AUC<sub>D</sub> – decremental area under the curve with respect to decrease

### Influence of chronic stress on basal BDNF

Chronic stress is a significant predictor of basal serum BNDF concentrations in both conditions (stress: *F*(1,27) = 4.320, Adj. R² =.14, *p* ≤.05; resting: *F*(1,27) = 4.483, Adj. R² =.14, *p* ≤.05). A higher chronic stress is associated with an increase in basal serum BDNF concentration and vice versa.

## 4. Discussion

We assessed the responses in BDNF and cortisol stress to a standardized stress task as well as their recovery. The applied stressor was sufficient to induce robust increases in BDNF and cortisol, which was not the case in the control condition. Interestingly, a strong cortisol response to stress was linked to an accelerated BDNF decline post-stress. Furthermore, higher chronic stress levels were linked to lower basal BDNF.

The stress-induced increases in BDNF and cortisol in our current study are well in line with earlier studies that also applied the TSST [14–16]. The activation of the acute stress system activation triggers a cascade of physiological changes, including the release of the stress hormone cortisol and the upregulation of BDNF. The latter likely serves as a neuroprotective mechanism to maintain brain health and cognitive function during stress [6].

The most important finding of our current study is the link between the cortisol response to stress and the subsequent BDNF recovery. This supports the hypothesis of Linz et al. [15] of an antagonistic relation between cortisol and BDNF in response to stress.

While animal models demonstrated an increase in both cortisol and hippocampal *Bdnf* mRNA expression in response to short-term stressors [38]. However, studies in animals furthermore demonstrated that elevated stress cortisol levels decrease hippocampal *Bdnf* mRNA expression [39,40]. This mechanistic data together with our findings and those from Linz et al. [15], highlight a dynamic relationship between cortisol and BDNF during stress. Initially, both factors increase in response to stress. Yet, as the stress response concludes, GC signaling could suppress BDNF. This pattern suggests an adaptive mechanism: an initial boost in BDNF could enhance cognitive function and neural plasticity during stress, while its subsequent suppression by GC signaling may help conserve energy and return the system to a baseline state once the stressor has passed [4,5,24].

In line with this hypothesis, we found high chronic stress perception to be linked to low basal serum BDNF levels. A comparable cross-sectional result of lower basal BDNF levels and higher psychological job stress was also observed in hospital employees [18]. In rodents, repeated restraint stress over 2-3 weeks reduced hippocampal *Bdnf* [17]. McEwen’s allostatic load model suggests that ongoing stress harms brain structures and overburdens the stress system, leading to prolonged high cortisol levels [1,21]. These repeated elevations in cortisol could decrease BDNF, affect neurogenesis, and contribute to neurodegenerative diseases [41]. Understanding the effects of chronic stress on BDNF may help to develop interventions and therapeutic strategies to mitigate the negative effects of chronic stress on the brain and overall well-being [42,43].

A major strength of this study is the within-subjects design using the standardized and reliable psychosocial stress test (TSST) and a resting control condition regarding to the circadian rhythm of BDNF [44]. In addition, there was a frequent assessment of serum BDNF and cortisol to accurately measure stress-induced dynamics, including peaks and recovery processes.

However, the present study has some limitations. Given the known age- and gender- specific differences in BDNF levels [27,28], we limited our study to young male participants. Thus, further research need to test our findings in women and older persons. Finally, in the present study serum BDNF levels were measured which may not necessarily fully reflect the regulation of BDNF in the hippocampus [45–47].

In conclusion, the present study showed that acute psychosocial stress increases serum BDNF and cortisol. The rise in cortisol appears to regulate the decline in BDNF post-stress. Chronic stress could lead to significant changes in BDNF in the circulation and presumably also in the brain. This could contribute to reduced neurogenesis, and an increased risk of neurodegenerative conditions in persons suffering from chronic stress. This could be the basis for interventions that can offset the detrimental effects of chronic stress on BDNF with possible long-term benefits for brain health and overall well-being.

## Funding

This research received no external funding.

## Data availability statement

Raw data produced in the present study are available upon reasonable request to the authors

## Ethics Statement

The studies involving human participants were reviewed and approved by Local Ethics Committee of the Landesärztekammer Rheinland-Pfalz. The participants provided their written informed consent to participate in this study.

## Funding

This research received no external funding.

## Acknowledgments

We thank the medical PhD students J.K., A.B., S.T., L.B., S.B. & E.K. for assisting with data collection.

## Conflicts of Interest

The authors declare no conflict of interest.

## Notes

### Competing Interest Statement

The authors have declared no competing interest.

### Author Declarations

The studies involving human participants were reviewed and approved by Local Ethics Committee of the Landesärztekammer Rheinland-Pfalz.

## References

1. McEwen BS. Neurobiological and Systemic Effects of Chronic Stress. Chronic Stress. 2017;1:247054701769232.

2. WHO. The World Health Report 2001. Mental Health: New Understanding, New Hope. WHO; 2001.

3. Chu B, Marwaha K, Sanvictores T, Ayers D. Physiology, Stress Reaction. StatPearls Publishing; 2021.

4. Egeland M, Zunszain PA, Pariante CM. Molecular mechanisms in the regulation of adult neurogenesis during stress. Nat Rev Neurosci. 2015;16:189–200.

5. Tsigos C, Chrousos GP. Hypothalamic–pituitary–adrenal axis, neuroendocrine factors and stress. J Psychosom Res. 2002;53:865–871.

6. Miranda M, Morici JF, Zanoni MB, Bekinschtein P. Brain-Derived Neurotrophic Factor: A Key Molecule for Memory in the Healthy and the Pathological Brain. Front Cell Neurosci. 2019;13.

7. Acheson A, Conover JC, Fandl JP, DeChiara TM, Russell M, Thadani A, et al. A BDNF autocrine loop in adult sensory neurons prevents cell death. Nature. 1995;374:450–453.

8. Zigova T, Pencea V, Wiegand SJ, Luskin MB. Intraventricular Administration of BDNF Increases the Number of Newly Generated Neurons in the Adult Olfactory Bulb. Molecular and Cellular Neuroscience. 1998;11:234–245.

9. Tapia-Arancibia L, Rage F, Givalois L, Arancibia S. Physiology of BDNF: focus on hypothalamic function. Front Neuroendocrinol. 2004;25:77–107.

10. Erickson KI, Prakash RS, Voss MW, Chaddock L, Heo S, McLaren M, et al. Brain-Derived Neurotrophic Factor Is Associated with Age-Related Decline in Hippocampal Volume. Journal of Neuroscience. 2010;30:5368–5375.

11. Wilson RS, Schneider JA, Boyle PA, Arnold SE, Tang Y, Bennett DA. Chronic distress and incidence of mild cognitive impairment. Neurology. 2007;68:2085–2092.

12. Lee B-H, Kim Y-K. The Roles of BDNF in the Pathophysiology of Major Depression and in Antidepressant Treatment. Psychiatry Investig. 2010;7:231.

13. Mori Y, Tsuji M, Oguchi T, Kasuga K, Kimura A, Futamura A, et al. Serum BDNF as a Potential Biomarker of Alzheimer’s Disease: Verification Through Assessment of Serum, Cerebrospinal Fluid, and Medial Temporal Lobe Atrophy. Front Neurol. 2021;12.

14. Meng D, Wu T, Rao U, North CS, Xiao H, Javors MA, et al. Serum NPY and BNDF response to a behavioral stressor in alcohol-dependent and healthy control participants. Psychopharmacology (Berl). 2011;218:59–67.

15. Linz R, Puhlmann LMC, Apostolakou F, Mantzou E, Papassotiriou I, Chrousos GP, et al. Acute psychosocial stress increases serum BDNF levels: an antagonistic relation to cortisol but no group differences after mental training. Neuropsychopharmacology. 2019;44:1797–1804.

16. Hermann R, Schaller A, Lay D, Bloch W, Albus C, Petrowski K. Effect of acute psychosocial stress on the brain-derived neurotrophic factor in humans – a randomized cross within trial. Stress. 2021;24:442–449.

17. Murakami S, Imbe H, Morikawa Y, Kubo C, Senba E. Chronic stress, as well as acute stress, reduces BDNF mRNA expression in the rat hippocampus but less robustly. Neurosci Res. 2005;53:129–139.

18. Mitoma M, Yoshimura R, Sugita A, Umene W, Hori H, Nakano H, et al. Stress at work alters serum brain-derived neurotrophic factor (BDNF) levels and plasma 3-methoxy-4- hydroxyphenylglycol (MHPG) levels in healthy volunteers: BDNF and MHPG as possible biological markers of mental stress? Prog Neuropsychopharmacol Biol Psychiatry. 2008;32:679–685.

19. Jeanneteau FD, Lambert WM, Ismaili N, Bath KG, Lee FS, Garabedian MJ, et al. BDNF and glucocorticoids regulate corticotrophin-releasing hormone (CRH) homeostasis in the hypothalamus. Proceedings of the National Academy of Sciences. 2012;109:1305–1310.

20. Castren E, Thoenen H, Lindholm D. Brain-derived neurotrophic factor messenger RNA is expressed in the septum, hypothalamus and in adrenergic brain stem nuclei of adult rat brain and is increased by osmotic stimulation in the paraventricular nucleus. Neuroscience. 1995;64:71–80.

21. McEwen BS. Protective and Damaging Effects of Stress Mediators. New England Journal of Medicine. 1998;338:171–179.

22. Herman JP, McKlveen JM, Ghosal S, Kopp B, Wulsin A, Makinson R, et al. Regulation of the Hypothalamic-Pituitary-Adrenocortical Stress Response. Compr Physiol, Wiley; 2016. p. 603–621.

23. Givalois L, Naert G, Rage F, Ixart G, Arancibia S, Tapia-Arancibia L. A single brain-derived neurotrophic factor injection modifies hypothalamo–pituitary–adrenocortical axis activity in adult male rats. Molecular and Cellular Neuroscience. 2004;27:280– 295.

24. Suri D, Vaidya VA. Glucocorticoid regulation of brain-derived neurotrophic factor: Relevance to hippocampal structural and functional plasticity. Neuroscience. 2013;239:196–213.

25. Wittchen HU, Zaudig M, Fydrich T. Strukturiertes Klinisches Interview für DSM-IV. Göttingen: Hogrefe; 1997.

26. APA. Diagnostic and Statistical Manual of Mental disorders. DSM-IV-TR. 4th ed. Washington DC: American Psychiatric Association; 2000.

27. Chan CB, Ye K. Sex differences in brain-derived neurotrophic factor signaling and functions. J Neurosci Res. 2017;95:328–335.

28. Oh H, Lewis DA, Sibille E. The Role of BDNF in Age-Dependent Changes of Excitatory and Inhibitory Synaptic Markers in the Human Prefrontal Cortex. Neuropsychopharmacology. 2016;41:3080–3091.

29. Kirschbaum C, Pirke KM, Hellhammer DH. The ‘Trier Social Stress Test’--a tool for investigating psychobiological stress responses in a laboratory setting. Neuropsychobiology. 1993;28:76–81.

30. Gaab J. PASA - Primary appraisal secondary appraisal. Ein Fragebogen zur Erfassung von situationsbezogenen kognitiven Bewertungen. Verhaltenstherapie. 2009;19:114–115.

31. Beck AT, Steer RA, Brown GK. Beck Depression Inventory: second edition manual. 2nd ed. San Antonio, TX: The Psychological Corporation; 1996.

32. Cohen S, Kamarck T, Mermelstein R. A Global Measure of Perceived Stress. J Health Soc Behav. 1983;24:385.

33. Derogatis L. Administration, scoring, and procedures manual for the SCL-90-R. Baltimore: Clinical Psychometric Research; 1977.

34. Schulz P, Schlotz W, Becker P. Trierer Inventar zum chronischen Stress (TICS). Göttingen: Hogrefe; 2004.

35. Faul F, Erdfelder E, Lang A-G, Buchner A. G*Power 3: A flexible statistical power analysis program for the social, behavioral, and biomedical sciences. Behav Res Methods. 2007;39:175–191.

36. Pruessner JC, Kirschbaum C, Meinlschmid G, Hellhammer DH. Two formulas for computation of the area under the curve represent measures of total hormone concentration versus time-dependent change. Psychoneuroendocrinology. 2003;28:916–931.

37. Fekedulegn DB, Andrew ME, Burchfiel CM, Violanti JM, Hartley TA, Charles LE, et al. Area Under the Curve and Other Summary Indicators of Repeated Waking Cortisol Measurements. Psychosom Med. 2007;69:651–659.

38. Neeley EW, Berger R, Koenig JI, Leonard S. Strain dependent effects of prenatal stress on gene expression in the rat hippocampus. Physiol Behav. 2011;104:334–339.

39. Lee T, Saruta J, Sasaguri K, Sato S, Tsukinoki K. Allowing animals to bite reverses the effects of immobilization stress on hippocampal neurotrophin expression. Brain Res. 2008;1195:43–49.

40. Hansson AC, Sommer WH, Metsis M, Stromberg I, Agnati LF, Fuxe K. Corticosterone Actions on the Hippocampal Brain-Derived Neurotrophic Factor Expression are Mediated by Exon IV Promoter. J Neuroendocrinol. 2006;18:104–114.

41. Numakawa T, Odaka H, Adachi N. Actions of Brain-Derived Neurotrophic Factor and Glucocorticoid Stress in Neurogenesis. Int J Mol Sci. 2017;18:2312.

42. Puhlmann L, Vrtička P, Linz R, Papassotiriou I, Chrousos GP, Engert V, et al. Serum BDNF increase and the role of cortisol reduction following contemplative mental training. Psychoneuroendocrinology. 2021;131:105510.

43. de Lima NS, De Sousa RAL, Amorim FT, Gripp F, Diniz e Magalhães CO, Henrique Pinto S, et al. Moderate-intensity continuous training and high-intensity interval training improve cognition, and BDNF levels of middle-aged overweight men. Metab Brain Dis. 2022;37:463–471.

44. Begliuomini S, Lenzi E, Ninni F, Casarosa E, Merlini S, Pluchino N, et al. Plasma brain-derived neurotrophic factor daily variations in men: correlation with cortisol circadian rhythm. Journal of Endocrinology. 2008;197:429–435.

45. Filimonova EA, Pashkov AA, Moysak GI, Tropynina AY, Zhanaeva SY, Shvaikovskaya AA, et al. Brain but not serum BDNF levels are associated with structural alterations in the hippocampal regions in patients with drug-resistant mesial temporal lobe epilepsy. Front Neurosci. 2023;17.

46. Klein AB, Williamson R, Santini MA, Clemmensen C, Ettrup A, Rios M, et al. Blood BDNF concentrations reflect brain-tissue BDNF levels across species. Int J Neuropsychopharmacol. 2011;14:347–353.

47. Puhlmann LMC, Linz R, Valk SL, Vrticka P, Vos de Wael R, Bernasconi A, et al. Association between hippocampal structure and serum Brain-Derived Neurotrophic Factor (BDNF) in healthy adults: A registered report. Neuroimage. 2021;236:118011.

